# ACCURACY OF COMPRESSED SENSING ACCELERATED 3D-FLAIR IN THE DIAGNOSTIC WORK-UP OF MULTIPLE SCLEROSIS AT 1.5 T

**DOI:** 10.64898/2026.11.02.26367062

**Authors:** Catalin George Iacoban, Claudia Klüner, Holger Bösenberg, Olga Alykova, Maike Gerdes, Sigrid Reuter, Christian Mathys, Benedikt Sundermann

## Abstract

**BACKGROUND:** 3D-FLAIR is central to MR-based diagnosis and monitoring of MS, but long acquisition times remain a limitation.

**OBJECTIVES:** This single-center prospective non-inferiority study evaluated whether compressed sensing (CS) – accelerated 3D-FLAIR at 1.5T provides lesion detection comparable to optimized Generalized Autocalibrating Partially Parallel Acquisition (GRAPPA) – accelerated 3D-FLAIR. Optimized CS- and GRAPPA-accelerated 3D-FLAIR sequences were incorporated into MRI examinations in patients with known or suspected MS. The primary endpoint was manual lesion count by two blinded readers using a 10% non-inferiority margin. Secondary endpoints included image quality, SNR, CNR, and automated lesion count and volumetry.

**RESULTS:** Sixty-eight patients were included (mean age, 40.1±13.3 years; 61.8% women). CS-accelerated 3D-FLAIR reduced acquisition time by 31.8% compared with GRAPPA (4:00 vs 5:52 minutes). Non-inferiority for manual lesion count was demonstrated for both readers (reader A, p < .0001; reader B, p = .005). Interrater agreement was excellent for CS (ICC = 0.921; 95% CI, 0.837-0.957) and very good for GRAPPA (ICC = 0.851; 95% CI, 0.769-0.906). Subjective image quality was high for both techniques. GRAPPA showed higher objective signal-to-noise-ratio (SNR) and contrast-to-noise-ratio (CNR), whereas automated lesion counts and volumes were higher for CS.

**CONCLUSION:** CS-accelerated 3D-FLAIR provided non-inferior lesion detection with substantial scan-time reduction.

## INTRODUCTION

Lengthy examinations and motion artifacts remain major limitations of MR.^1,2^ Reducing scan time may improve patient comfort and decrease motion-related artifacts. To address these challenges, advanced acceleration techniques are now widely available on recent clinical scanner generations.^3,4^

Parallel imaging (PI) methods such as Generalized Autocalibrating Partially Parallel Acquisition (GRAPPA) and Controlled Aliasing in Parallel Imaging Results in Higher Acceleration (CAIPIRINHA) have reduced scan times across various MRI sequences.^5–8^ More recently, CS has emerged as an additional acceleration strategy based on undersampled MR data acquisition and iterative reconstruction, enabling reduced scan times or improved spatial resolution.^9^ However, achievable acceleration with CS is limited by potential image quality degradation and the occurrence of reconstruction-related artifacts.^10,11^ MRI is the gold standard in the diagnostic work-up of MS, with high-resolution isotropic 3D-FLAIR serving as a key sequence for lesion detection and disease monitoring.^12,13^ Owing to its intrinsic sparsity and long acquisition time, 3D-FLAIR is particularly well suited for CS-based acceleration.^9^ Prior studies have shown that compressed-SENSE accelerated 3D-FLAIR reduces scan time while maintaining image quality and may additionally allow increased spatial resolution in routine clinical brain MRI.^14–16^ CS-accelerated 3D-FLAIR in MS cohorts has mainly been studied at 3T, demonstrating substantial scan-time reduction with preserved diagnostic image quality.^17^

Most studies were conducted at 3T and rely on vendor-specific (PHILIPS) compressed-SENSE implementations, limiting generalizability. Evidence comparing CS with established PI techniques such as GRAPPA or CAIPIRINHA at 1.5T remains scarce, and, to our knowledge, CS-accelerated 3D-FLAIR for MS lesion assessment has not yet been systematically evaluated at 1.5T in direct comparison with conventional PI methods. This prospective noninferiority study compared optimized CS-and GRAPPA-accelerated 3D-FLAIR in clinical routine, using manual lesion count as the primary endpoint and aspects of image quality as well as automated lesion count and volume as exploratory secondary endpoints. We followed the STARD and CONSORT reporting guidelines.

## MATERIALS AND METHODS

### RESEARCH DESIGN

This single-center prospective non-inferiority study was approved by the institutional ethics committee of Carl von Ossietzky Universität, Oldenburg, Niedersachsen, Germany (registration no. 2021-033). Sample size was estimated from a pilot retrospective analysis of routine clinical MRI data. Written informed consent was obtained from all participants in accordance with the Declaration of Helsinki. No contrast agent or sedative medication was administered for study purposes. However, patients who received contrast agents and/or sedation for clinical reasons were eligible.

During the first stage, sequence parameters for newer (CS, CAIPIRINHA) and conventional (GRAPPA) acceleration techniques were optimized in five healthy participants using a pragmatic empirical approach (see supplementary material). Based on expert consensus, the best-performing sequence from each acceleration arm was selected for the clinical phase (Table 1).

**Table 1.** Imaging parameters for the selected CS-and GRAPPA-accelerated 3D-FLAIR.

| **Parameters** | **CS-3D-FLAIR** | **GRAPPA-3D-FLAIR** |
|---|---|---|
| TE/TR/TI (ms) | 386,00 / 7000,0 / 2050 | 385,00 / 7000,0 / 2050 |
| FA (Flip Angle) | Variable | Variable |
| FOV (Frequency x Phase) (mm) | 250 x 250 | 250 x 250 |
| Slice Thickness (mm) | 1,0 mm | 1,0 mm |
| Matrix (Frequency x Phase) | 256 x 256 | 256 x 205 |
| Bandwidth (Hz/Pixel) | 698 | 651 |
| CS Acceleration Factor | 6.4 | N/A |
| GRAPPA Factor | N/A | 4 |
| Reconstruction Interpolation | 20% | 20% |
| Scan time (min:sec) | 4:00 | 5:52 |
| Denoising Factor | 10% | 10% |

During the second stage, the previously optimized best CS-accelerated and GRAPPA-accelerated sequences were added to routine clinical MRI examinations of patients with known or high clinical suspicion of MS, as indicated by neurological referral information. Both sequences were acquired natively, although the overall protocol included contrast administration. The acquisition order of CS and GRAPPA was systematically alternated. The subsequent comparison included an assessment of general and specific aspects of image quality as well as lesion load, counted both manually and automatically.

### ENDPOINTS

The primary endpoint was manual lesion count, with the primary hypothesis that optimized CS-accelerated FLAIR would not result in a clinically relevant reduction in counted lesions (>10%) compared with GRAPPA-accelerated FLAIR. Secondary endpoints included comparison of overall image quality (numerical rating scale), general image artifacts (numerical rating scale), motion artifacts (numerical rating scale), SNR and CNR (numerical rating scale and ROI-based measurements), lesion delineation (numerical rating scale), as well as automatic lesion count and volumetry.

### PATIENT COHORT

During sequence optimization, 5 healthy volunteers (≥18 years) were recruited. Based on the sample size estimation pilot study, the final cohort comprised 72 patients with known or suspected MS and at least two FLAIR-hyperintense lesions. Between July 2024 and February 2025, 72 consecutive patients at the Institute of Radiology and Neuroradiology, Evangelisches Krankenhaus Oldenburg, underwent MRI at 1.5T as clinically indicated. Standard MRI contraindications and specific CNS disorders were exclusion criteria (see supplementary material).

### IMAGING PROTOCOL

Imaging was performed on our clinical 1.5 T whole-body MR scanner (Magnetom Sola 1.5T; SIEMENS Healthineers, Erlangen, Germany; Software version XA20, 20 channel head/neck receiver coil, installed February 2020). Primary image data and technical parameters were stored in standard DICOM format.

The individualized MRI protocol included the two optimized CS-and GRAPPA-accelerated 3D-FLAIR sequences in addition to clinically indicated sequences.

A CS acceleration factor of 6.4 was applied, resulting in a scan time of 4:00 min:s. For GRAPPA-accelerated FLAIR, a GRAPPA factor of 4 was used, resulting in a scan time of 5:52 min:s (31.8% scan time reduction). Both 3D-FLAIR datasets were acquired in the sagittal plane. Detailed imaging parameters are summarized in Table 1.

### IMAGE ANALYSIS

All 3D-FLAIR datasets were pseudonymized, randomized, and independently assessed by two blinded readers (7 (CGI) and 14 years’ (BS) neuroradiology experience). Subjective image quality parameters were rated on a 100-point scale (0 = poor (insufficient for diagnosis / severe artifacts), 50 = adequate (moderate artifacts not impairing diagnosis), 100 = excellent (no artifacts)) across predefined criteria (see endpoints). Visible FLAIR-hyperintense lesions were manually counted in all locations. An isolated lesion was defined as a focal T2-hyperintensity surrounded by normal isointense white matter. Confluent lesions were counted as single lesions. The number of detected lesions was recorded as a numerical integer value. Additionally, automated lesion counting and volumetry was performed using the open-source Lesion Segmentation Toolbox (LST; https://www.applied-statistics.de/lst.html) for SPM (version 3.0.0, RRID:SCR_007037). All lesions, including those in deep white matter, were segmented using the lesion prediction algorithm proposed by Schmidt.^18^ SNR and CNR were calculated for each FLAIR acquisition using predefined ROIs in lesions, normal appearing white matter, and background noise (see supplementary material).

### STATISTICAL ANALYSIS

Statistical analyses were performed using SPSS Version 30 (IBM, Armonk, NY, USA) and SAS^®^ Analytics Software Online Studio (SAS Institute, Cary, NC, USA). Standard descriptive statistics were applied.

Non-inferiority of CS relative to GRAPPA for the primary endpoint (manual lesion count) was assessed separately for each rater using log-transformed scores. This was done using a one-sided t test with a prespecified non-inferiority margin of 10%, a difference considered clinically irrelevant for overall diagnostic performance.^19^ Results were visualized using box- and-whisker plots. Non-inferiority was concluded if the lower bound of the one-sided 95% confidence interval for the mean difference (CS-GRAPPA) exceeded the predefined margin. Statistical significance was set at *p* < 0.05.

Secondary exploratory endpoints were summarized for each variable as median (interquartile range). Visual comparisons of secondary endpoints were performed using split-violin plots to explore potential determinants of the main result (primary endpoint).

Interrater agreement between rater A and rater B for both CS and GRAPPA for all endpoints was reported as single-measure intraclass correlation coefficient derived from a two-way mixed-effects model with absolute agreement.

## RESULTS

### COHORT CHARACTERISTICS

Initially, 72 patients with known or clinically suspected MS were recruited and provided written informed consent. One patient was excluded because he had been enrolled twice during a follow-up examination. Two patients did not undergo MRI because the examination was cancelled by the treating physicians after consent had been obtained, and one patient was excluded due to severe motion artifacts. This resulted in a final study cohort of 68 patients. The mean age of participants was 40.1 ± 13.3 years, and 42 patients (61.8%) were female. The mean body mass index (BMI) was 26.1 ± 5.89 kg/m². Most patients were diagnosed with MS (n = 36, 52.9%), whereas in 32 patients (47.1%) the initial clinical diagnostic suspicion was not confirmed after a mean follow-up period of seven months.

### PRIMARY ENDPOINT

CS accelerated-FLAIR was non-inferior to GRAPPA-accelerated FLAIR, with no greater than 10 % reduction in manual lesion count for both Rater A *p* < .0001 and Rater B *p* = .005 (Figure 1). Interrater agreement for lesion count was excellent for CS-accelerated FLAIR and very good for GRAPPA-accelerated FLAIR (Table 2).

**Fig 1.**
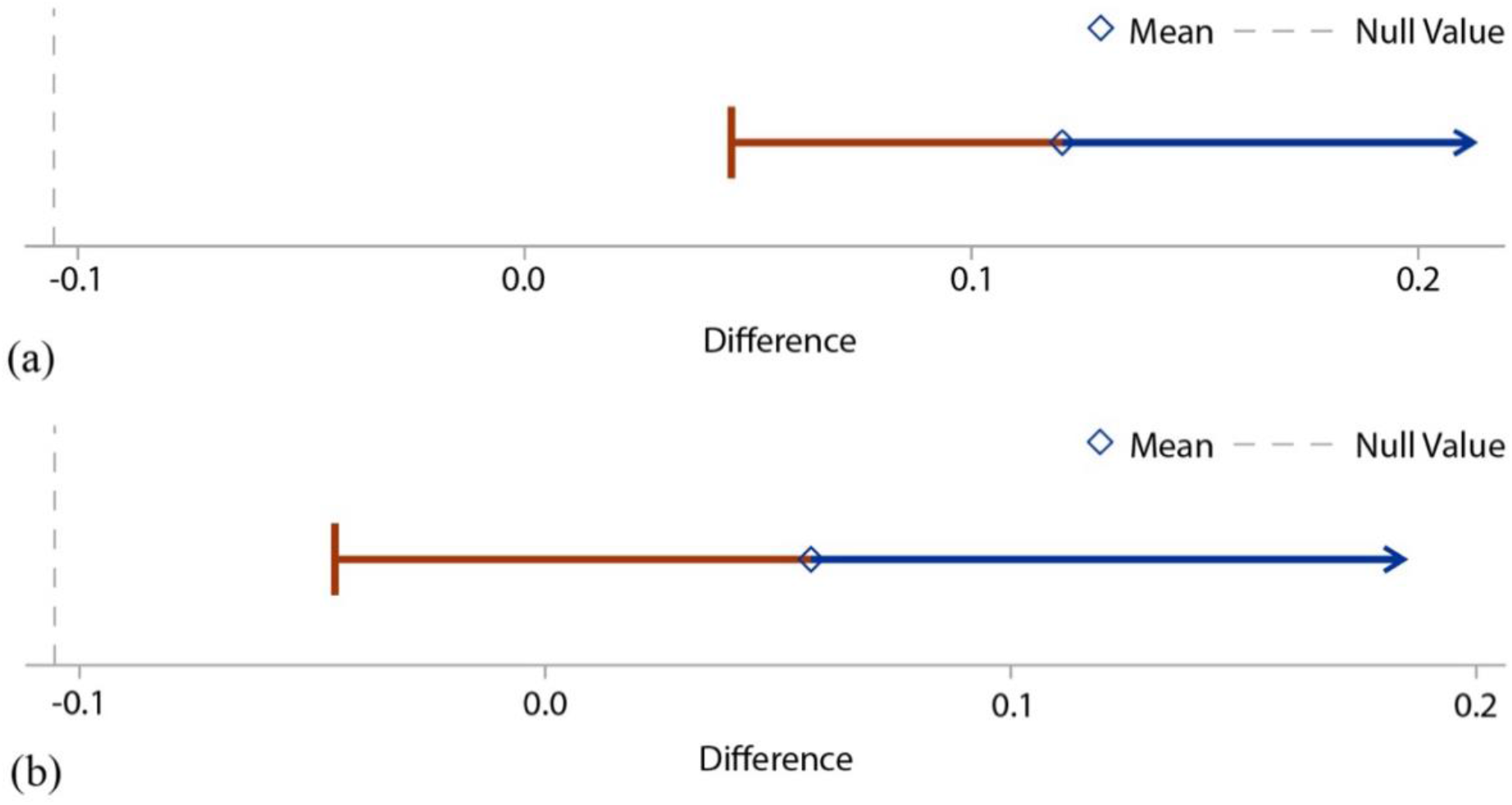
Results of the non-inferiority analysis of lesion counts for Rater A (a) and Rater B (b). The vertical dashed line on the far left indicates the non-inferiority margin (10%) obtained after logarithmic transformation of the mean differences between the two sequences. The diamond on the horizontal arrow represents the mean difference. Inferiority was rejected for both Rater A and Rater B, as the lower confidence limits (left-sided whiskers of the one-sided 95% confidence interval) lay well above the non-inferiority margin

**Table 2.** Interrater agreement, manual lesion counts, and automated lesion metrics for CS- and GRAPPA-accelerated FLAIR.

| **Lesion Count** | **Rater A** | **Rater B** | **ICC** | **95% CI** | **p** | **Auto Lesion Count** | **Auto Lesion Volume** |
|---|---|---|---|---|---|---|---|
| **CS – FLAIR** | 23.5 (11, 45.5) | 20.5 (9, 33.5) | .921 | (.837, .957) | <.001 | 12 (7.5,20) | 1.24 (.35, 3.41) |
| **GRAPPA – FLAIR** | 19.5 (8, 47) | 22 (8, 41) | .851 | (.769, .906) | <.001 | 8 (3.5, 16.5) | .66 (.16, 2.26) |

ICC = intraclass correlation coefficient; CI = confidence interval. p-values and CIs refer to ICC estimates.

### SECONDARY ENDPOINTS

Subjective image quality ratings for CS-and GRAPPA-accelerated FLAIR are summarized in Table 3 and Figures 2 and 3. Median scores were high across all subjective parameters for both acceleration techniques, with substantial overlap in score distributions between methods and raters. Rater A did not show a significant preference between methods, except for motion artifacts, which were rated slightly poorer with GRAPPA than CS. In contrast, Rater B showed a slight preference for CS across all subjective image quality measures. Inter-rater agreement ranged from moderate to good across metrics, with higher agreement for GRAPPA-accelerated FLAIR, particularly for overall image quality and lesion detection. Because these secondary endpoints are exploratory, results were summarized descriptively with assessment of interrater agreement and no non-inferiority testing was performed.

**Fig 2.**
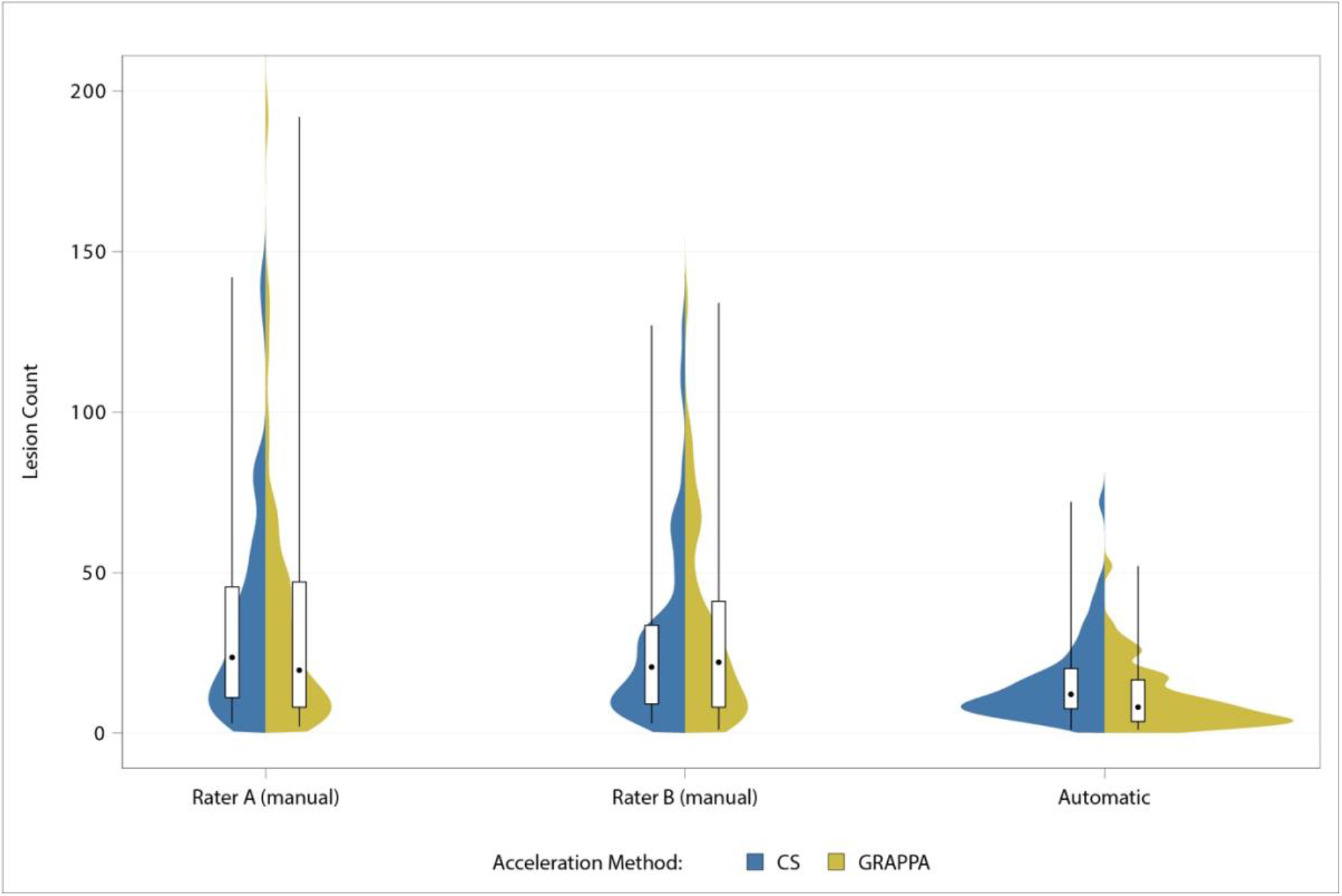
Distribution of manual and automated lesion counts by acceleration method

**Fig 3.**
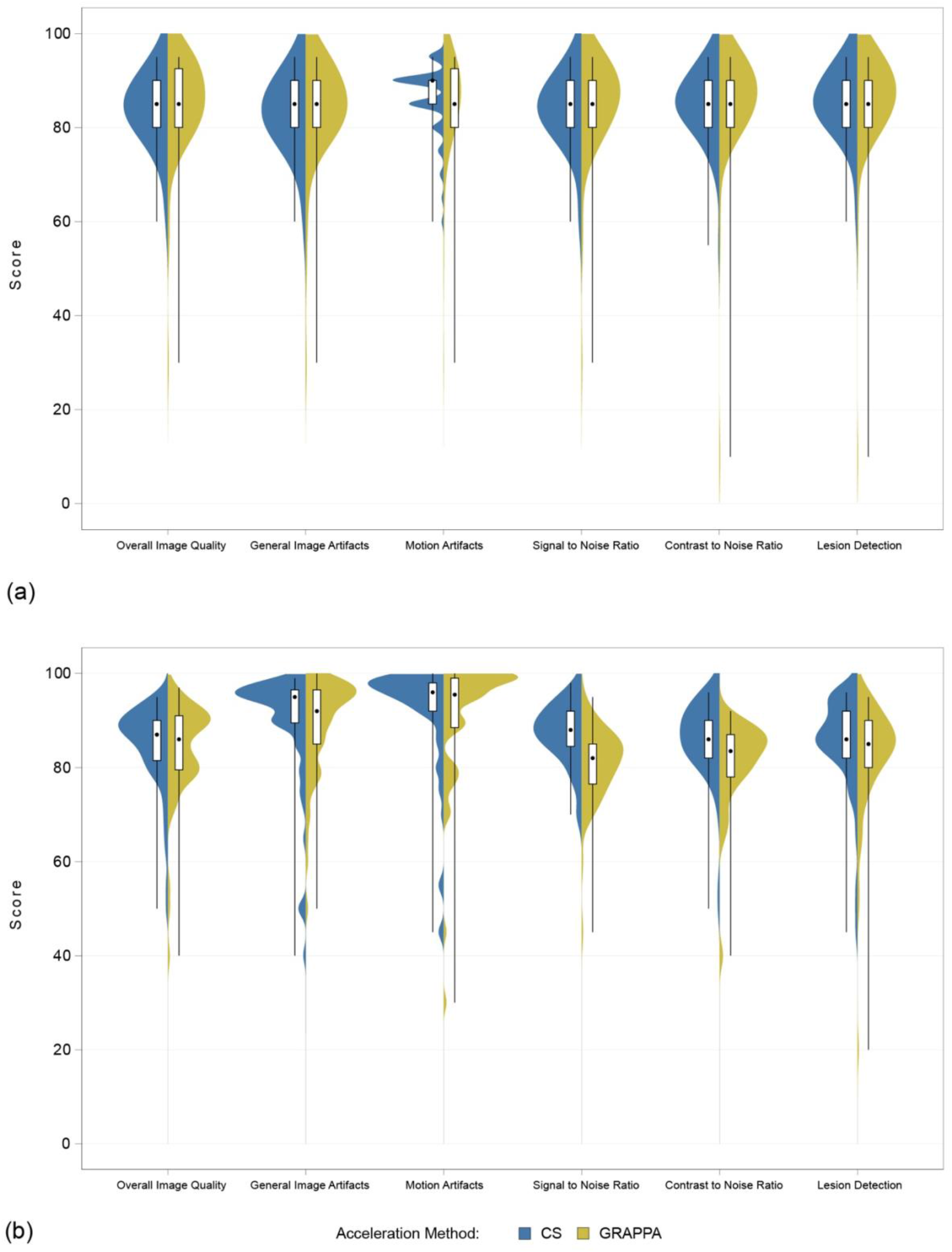
Subjective Image Quality Assessments by FLAIR acceleration Method in Rater A (a) and Rater B (b). The serrated appearance of Rater A’s CS motion artifacts half-violin results from scores being assigned in 5-point increments on the 0-100 scale

**Table 3.** Subjective image quality ratings by sequence acceleration type and rater (0-100 scale)

|  | **CS** |  |  |  |  | **GRAPPA** |  |  |  |  |
|---|---|---|---|---|---|---|---|---|---|---|
| **Parameter** | **Rater A** | **Rater B** | **ICC** | **95% CI (ICC)** | **p** | **Rater A** | **Rater B** | **ICC** | **95% CI (ICC)** | **p** |
| **Overall Image Quality** | 85 (80-90) | 87 (81.5-90) | .691 | (.544, .791) | <.001 | 85 (80-92.5) | 86 (79.5-91) | .839 | (.752, .898) | <.001 |
| **General Image Artifacts** | 85 (80-90) | 95 (89.5-96.5) | .548 | (.201, .743) | <.001 | 85 (80-90) | 92 (85-96.5) | .695 | (.326, .847) | <.001 |
| **Motion Artifacts** | 90 (85-90) | 96 (92-98) | .593 | (.273, .769) | <.001 | 85 (80-92.5) | 95.5(88.5-99) | .709 | (.285, .863) | <.001 |
| **Signal to Noise Ratio (subjective)** | 85 (80-90) | 88 (84.5-92) | .600 | (.281, .773) | <.001 | 85 (80-90) | 82 (76.5-85) | .547 | (.284, .718) | <.001 |
| **Contrast to Noise Ratio (subjective)** | 85 (80-90) | 86 (82-90) | .495 | (.294, .655) | <.001 | 85 (80-90) | 83.5 (78-87) | .683 | (.403, .824) | <.001 |
| **Lesion Detection** | 85 (80-90) | 86 (82-92) | .507 | (.306, .665) | <.001 | 85 (80-90) | 85 (80-90) | .797 | (.631, .883) | <.001 |

ICC = intraclass correlation coefficient; CI = confidence interval. p-values and CIs refer to ICC estimates.

ROI-based measurements demonstrated higher SNR and CNR for GRAPPA-accelerated FLAIR compared with CS-accelerated FLAIR, with median SNR values of 211.8 (189.6-247.3) versus 147.2 (123.6-169.0) and median CNR values of 140.3 (105.4-176.1) versus 124.9 (99.9-150.4), respectively.

**Fig 4.**
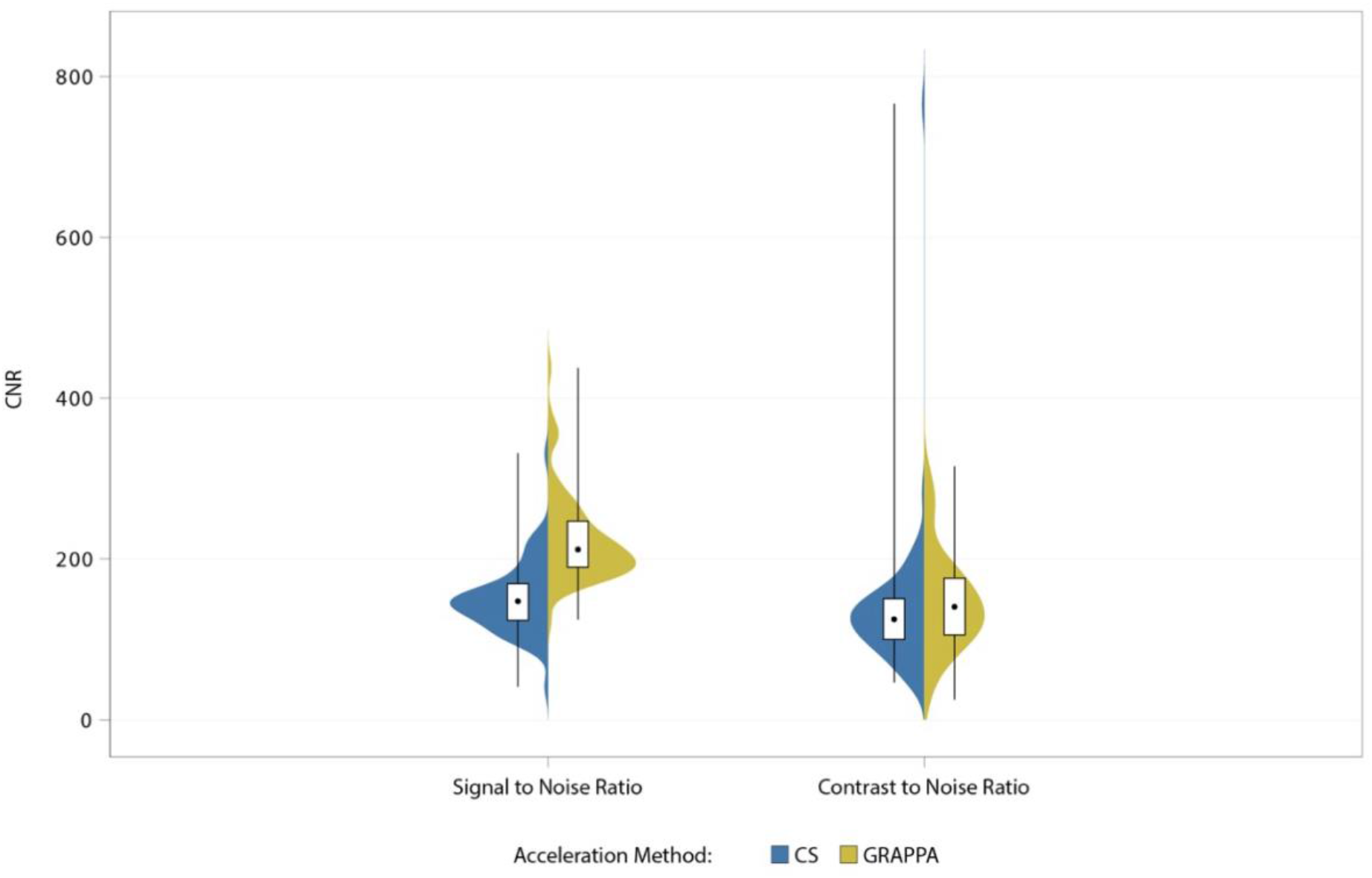
Distribution of ROI-based signal-to-noise and contrast-to-noise ratios by acceleration method

Automated lesion counts were lower for GRAPPA-accelerated FLAIR than for CS-accelerated FLAIR (Figure 2, Table 2). Similarly, automated lesion volume was lower for GRAPPA than for CS (Figure 5, Table 2).

**Fig 5.**
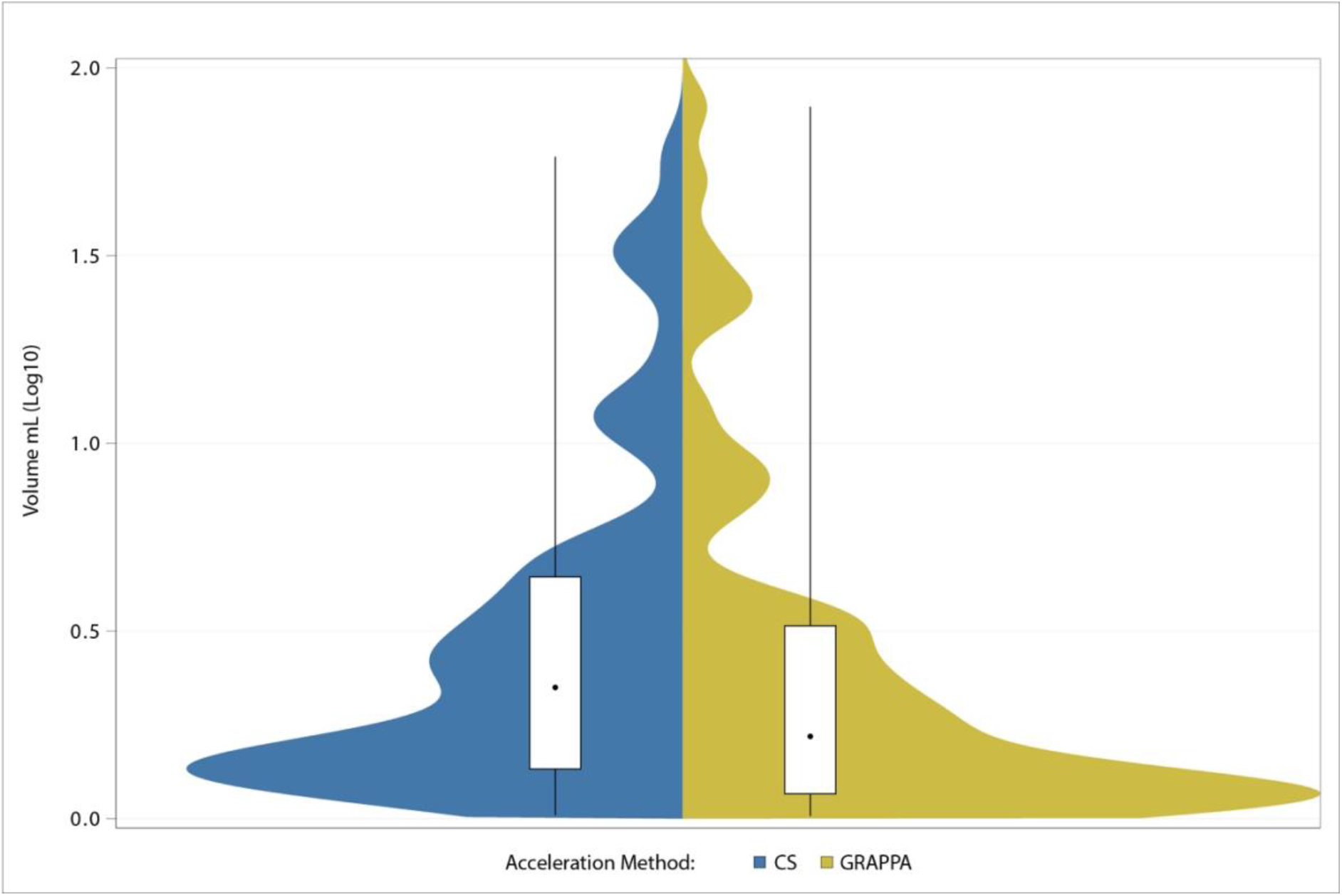
Automatic lesion volume (Log10 scale) by acceleration method

## DISCUSSION

MS diagnosis and treatment escalation decisions rely on frequent MR monitoring, with current MS imaging guidelines recommending 3D-FLAIR in the initial diagnostic work-up and during follow-up.^12,13^

Several acceleration techniques have emerged to speed brain imaging, reducing motion artifacts and scan times, particularly important for patients requiring frequent follow-up, such as those with MS.^8,9,20^ CS and parallel imaging techniques can significantly reduce MR scan times without compromising diagnostic performance, across various clinical applications, including neuroradiological.^14–17^

Prior studies support the feasibility of CS-acceleration in neuroimaging, despite variability in machine vendors, field strengths, and acceleration factors. However, the optimal balance among acceleration factor, scan-time reduction, and image quality remains debated.^14,16,17^ Molnár et al.^14^, Vranic et al.^15^ and Mönch et al.^16^ demonstrated that CS-accelerated 3D-FLAIR substantially reduces scan-time while maintaining image quality comparable to conventional acquisitions, with Mönch et al. additionally reporting improved spatial resolution.^16^ However, Vranic and Molnár used PHILIPS vendor-specific compressed-SENSE implementations at 3T, whereas we evaluated a SIEMENS CS approach at 1.5T, limiting direct comparability.

The most comparable study, by Toledano-Massiah et al., compared CS-accelerated with non-accelerated 3D-FLAIR in 23 patients with relapsing-remitting MS at 3T, demonstrating a 27% scan-time reduction with comparable image quality (SNR, CNR), lesion count, and automated lesion volume using a CS acceleration factor of 1.3.^17^

We evaluated whether faster CS-accelerated 3D-FLAIR (factor 6.4; 4:00 min) preserved lesion detection relative to GRAPPA-accelerated 3D-FLAIR (factor 4; 5:52 min) in patients with known or suspected MS at 1.5T. CS achieved non-inferior manual lesion counts with excellent interrater agreement, corresponding to a 31.8% scan-time reduction without clinically meaningful loss in lesion detection, consistent with prior evidence.^17^

Our study extends prior work by comparing CS with an optimized GRAPPA-accelerated reference, evaluating performance at the more widely available 1.5T, applying a prespecified non-inferiority margin to a clinically meaningful endpoint (manual lesion count), using a granular 0-100 image quality scale to capture subtle differences and including a larger cohort. Although only 52.9% of participants had confirmed MS, all had lesions.

Subjective image quality parameters showed mild reader-dependent differences, with Rater B favoring CS and Rater A showing no clear preference except for slightly worse GRAPPA motion artifacts. Moderate-to-good interrater agreement suggests these differences reflect individual preference rather than systematic differences between techniques.^21^

Although ROI-based measurements showed higher SNR and CNR for GRAPPA-accelerated FLAIR, subjective ratings by Rater B paradoxically favored CS. The difference between ROI-based SNR/CNR measurements in CS and GRAPPA may reflect differences in acceleration method reconstruction techniques. CS relies on undersampled k-space acquisition, which reduces measured signal and increases effective noise. However, the nonlinear iterative reconstruction suppresses incoherent noise, producing smoother background signal. These effects are particularly pronounced in sequences such as 3D-FLAIR. In contrast, GRAPPA utilizes coil sensitivity information and reconstructs missing k-space data, preserving signal and generally yielding higher SNR and CNR.^9,22,23^ Thus, the lower objective SNR/CNR values in CS may primarily reflect signal reduction rather than increased noise because its reconstruction suppresses low-intensity signals while denoising. This creates visually “cleaner” images that may receive higher subjective ratings despite lower quantitative metrics, potentially explaining Rater B’s preference. The interrater discrepancy may reflect different perceptual sensitivities to image texture, noise, or overall image impression. Therefore, subjective ratings do not necessarily align with purely mathematical relationships. Rater B might have favored the smooth, denoised CS appearance, while Rater A equally weighted both techniques, illustrating that subjective ratings do not necessarily align with quantitative metrics.^21^

Moreover, other MRI parameters might influence SNR and CNR. First, SNR is proportional to voxel volume and inversely related to the square root of receiver bandwidth. Therefore, smaller voxels and higher bandwidth reduce measured SNR even when diagnostic performance remains acceptable. This may partly explain the discrepancy, as our CS and GRAPPA sequences differed not only in acceleration method and factor but also in matrix and bandwidth, which can influence SNR and CNR independently of acceleration strategy.^24,25^ Second, quantitative noise and SNR/CNR measurements can be challenging when reconstruction extends beyond a simple Fourier transform. Parallel imaging reconstructions (including GRAPPA) introduce spatially varying noise amplification, described by the coil geometry factor (g-factor), complicating conventional noise ROI measurements and making SNR comparisons sensitive to ROIs placement.^26,27^ Similarly, CS reconstructions are nonlinear and can alter the appearance and distribution of residual artifacts, often producing noise-like texture under incoherent sampling. Consequently, the relationship between perceived image quality and measured SNR less straightforward than in fully sampled imaging.^28–31^

Automatic lesion counts and volumes were slightly higher in CS than GRAPPA, despite the lower SNR and CNR of CS acquisitions. However, lesion conspicuity depends not only on global SNR and CNR but also on image texture, background noise distribution, and edge definition. CS employs nonlinear iterative reconstruction with sparsity constraints, suppressing incoherent noise and smoothing homogeneous tissue while preserving structural interfaces. This may improve lesion separation, increasing lesion detection.^9,11^ Consequently, automated segmentation algorithms may identify lesions in CS images undetected in GRAPPA acquisitions, suggesting greater detection sensitivity. However, this increased sensitivity may potentially come at the expense of specificity, as noise suppression and altered image texture may enhance subtle signal variations and cause them to be misclassified as lesions. These may affect future AI-based image interpretation, as reconstruction techniques could influence automated detection performance. Therefore, reconstruction methods should be considered when developing and validating automated lesion detection tools.

Automated MS lesion quantification tools are not yet standardized, and few studies have compared automated outputs across acceleration methods or with manual lesion counts. The clinical relevance of lesions smaller than 3 mm remains debated, with the McDonald criteria considering only lesions of at least 3 mm in diameter.^12^ The discrepancy between manual and automated lesion counts may partly stem from this issue, as manual assessment included lesions regardless of size, whereas automated segmentation and volumetric tools may be less sensitive to very small lesions. We found higher automated lesion counts and higher volumes in CS-FLAIR than GRAPPA-FLAIR despite equivalent manual counts, suggesting reconstruction-dependent intensity distributions affect thresholding algorithms. Whether automated lesion quantification tools can reliably replace manual counting using different acceleration methods remains unresolved and warrants caution, particularly in longitudinal assessments using varying acquisition techniques. While automated approaches may offer a time-efficient complement to manual lesion counting, further research is needed to clarify method-specific differences, such as the types of lesions that may be missed by automatic volumetry in GRAPPA-acceleration or additionally detected in CS-acceleration. In our study, CS-based volumetry identified more small and isolated lesions (Figure S1, supplementary material), although the clinical relevance of these differences requires further investigation. Toledano-Massiah et al. found no automated volume differences between CS- and conventional FLAIR at 3T, contradicting our findings,^17^ potentially attributable to the different reconstruction families: CS vs. GRAPPA as opposed to CS vs. conventional FLAIR, different field strengths (1.5 T vs. 3 T) and to the higher acceleration factors we employed in CS (6.4 vs. 1.3 in their study), with higher CS factors introducing greater intensity distribution changes and apparent lesion boundaries, factors known to influence artifact profile and affect automated thresholding.

Several limitations should be acknowledged. First, only 52.9% of patients had confirmed MS, which may limit generalizability to lesion-rich patients. However, all participants had at least two lesions, supporting applicability to patients with clinical MS-suspicion. Second, the primary endpoint pooled all lesions rather than assessing lesion detection by anatomical location. Prior MS imaging studies stratified lesions by location, and the absence of this analysis may have obscured regional differences between sequences.^17^

Future studies could assess the robustness of automated MS lesion detection across different acceleration techniques and vendors, particularly in longitudinal settings, where technical variability may influence perceived disease progression and treatment decisions. AI- and deep learning-enhanced reconstruction combined with CS may further reduce scan time while preserving lesion detectability.^32–34^ Ultrafast EPI-FLAIR also warrants further investigation for routine MS monitoring, alone or in combination with CS.

## CONCLUSION

CS-accelerated 3D-FLAIR achieved noninferior lesion detection compared to GRAPPA with excellent interrater agreement and 31.8% shorter scans. Despite higher objective GRAPPA SNR and CNR, subjective image quality was high for both techniques. Automated volumetry yielded higher CS lesion counts and volumes, suggesting increased detection of small or isolated lesions, although their clinical relevance in longitudinal monitoring remains uncertain. Overall, CS-accelerated 3D-FLAIR at 1.5T is a time-efficient, robust GRAPPA alternative.

## Data Availability

The datasets generated and/or analyzed during the current study are not made publicly available due to local and European legal requirements governing the protection of patient data.

## ACKNOWLEDGEMENTS

We acknowledge the contribution of Dipl.-Statistikerin Nadine Bonberg, who assisted with the estimation of the sample size required for the study. ChatGPT (OpenAI) was used to assist with language editing and to improve the readability of the manuscript. All scientific content, interpretations, and conclusions were generated, reviewed, and verified entirely by the authors.

## STATEMENTS AND DECLARATIONS

### ETHICAL CONSIDERATIONS

This single-center prospective non-inferiority study was approved by the institutional ethics committee of Carl von Ossietzky Universität, Oldenburg, Germany (No. 2021-033).

### CONSENT TO PARTICIPATE

All participants provided written informed consent, and procedures complied with the Declaration of Helsinki and applicable regulations.

### CONSENT FOR PUBLICATION

The authors affirm that human research participant provided informed consent for publication of the images in Figure S1.

### AUTHORS’ CONTRIBUTIONS

Catalin George Iacoban, Christian Mathys and Benedikt Sundermann contributed to the initial study conception and design. Material preparation, data collection, and analysis were performed by Catalin George Iacoban, Claudia Klüner, Holger Bösenberg, Olga Alykova, Maike Gerdes, Sigrid Reuter, Christian Mathys, and Benedikt Sundermann. The first draft of the manuscript was written by Catalin George Iacoban, and all authors reviewed and commented on previous versions of the manuscript. All authors have read and approved the final manuscript.

### PRESENTATION STATEMENT

The results of this study were presented in poster format at the 49th Annual Meeting of the European Society of Neuroradiology (ESNR), held jointly with the XXIII Symposium Neuroradiologicum and the British Society of Neuroradiologists (BSNR) Annual Meeting, in London, United Kingdom, from 30 September to 4 October 2026, are part of the doctoral thesis of the first author and were submitted as a separate manuscript to a profile-specific medical journal.

