## supplementary material for "ACCURACY OF COMPRESSED SENSING ACCELERATED 3D-FLAIR IN THE DIAGNOSTIC WORK-UP OF MULTIPLE SCLEROSIS AT 1.5 T"

### PRECURSOR PILOT STUDY FOR SAMPLE SIZE ESTIMATION

Sample size was estimated based on a precursor pilot study with a retrospective analysis of routine clinical MRI data. The precursor pilot study, used for sample size estimation, involved retrospective analysis of fully anonymized medical images. Therefore, patient information and informed consent were waived.

Fully anonymized 3D-FLAIR sequences comparable to the planned study protocol, acquired between January 1 and December 31, 2021, were retrospectively retrieved from the PACS at the Institute of Radiology and Neuroradiology, Evangelisches Krankenhaus Oldenburg. The dataset included 20 chronologically consecutive routine MR examinations performed on the study scanner (1.5. T Magnetom Sola; SIEMENS Healthineers, Erlangen, Germany, Software version XA20, using a 20-channel head/neck receiver coil). MS lesions were manually counted, followed by statistical analysis of data distribution. Due to lognormal data distribution, the required sample size was calculated using a paired t-test for mean ratios. This method, available through SAS® Analytics Software, considers fixed parameters such as alpha error, nominal power, and inter-sequence correlations, while variations in results stem from differing coefficients of variation (CV). Based on these analyses, the estimated sample size was approximately 72 subjects, including 20% drop-out reserve (the 20% potential data exclusions due to motion artifacts or incomplete examinations).

### SEQUENCE OPTIMIZATION

During the first stage of the prospective study, sequence parameters for newer acceleration methods (CS, CAIPIRINHA) and for conventional acceleration techniques derived from parallel imaging (GRAPPA) were optimized in five healthy participants using an approach similar to routine clinical protocol optimization by radiologists or MRI technologists. This pragmatic strategy relied on empirical optimization within predefined boundaries rather than exhaustive grid searches or simulation-based optimization.

To optimize and select the imaging sequences, multiple 3D-FLAIR acquisitions with modified parameters (e.g., with and without CS, GRAPPA, and CAIPIRINHA acceleration) were performed sequentially in five healthy volunteers. Parameter selection was conducted through a targeted conjoint search by both a junior user (CGI) and an experienced user (BS). To identify the most suitable 3D-FLAIR sequences, key parameters were systematically varied within predefined guideline ranges. Total scan time was limited to a maximum of 60 minutes per volunteer examination. Acceleration factors were constrained as follows: GRAPPA acceleration factor 1-4, CAIPIRINHA acceleration factor 4-6, and compressed sensing (CS) acceleration factor 2-25. The number of averages ranged from 1 to 2. In contrast, nominal resolution and parameters determining image contrast were kept constant, including repetition time (TR), echo time (TE), and inversion time (TI). In total, 22 acquisitions were performed sequentially (see Diagram 1). Denoising levels were evaluated separately as a post-processing step. Scan time reductions achieved with each acceleration technique were quantified.

Sequence selection for further evaluation was based on consensus among experienced neuroradiologists (CM, BS, HB) and a general radiologist with 7 years' experience in neuroradiology (CGI). For each acceleration method arm (newer and conventional), the two best-performing sequences were independently assessed (CK, CGI) based on the overall image quality (using an image rating reference catalogue) and ultimately the optimal sequence per arm was selected for use in the subsequent clinical phase (Table 1).

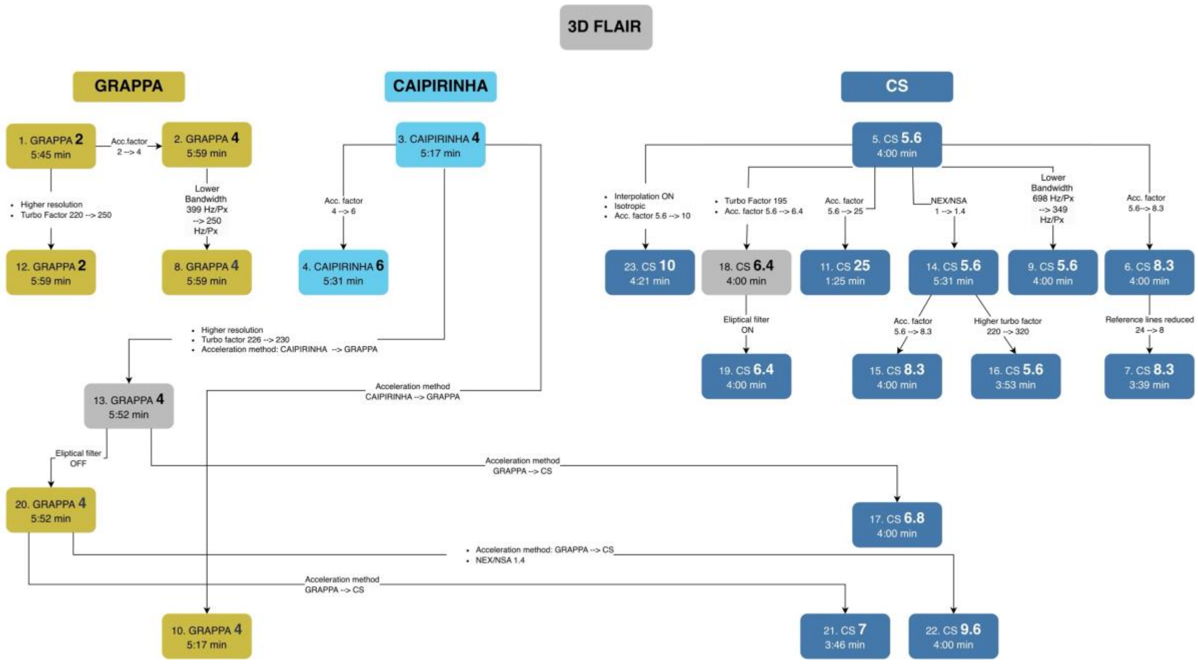


**Diagram S1.** Study workflow for the optimization and selection of 3D-FLAIR sequence parameters in healthy volunteers. A total 22 GRAPPA-, CAIPIRINHA-, and CS-accelerated sequences were performed sequentially in healthy volunteers. Sequences 13 (GRAPPA) and 18 (CS), highlighted in grey, represent the final optimized sequences selected for the subsequent phase of the study.

**PATIENT COHORT**

During the sequence-optimization stage, 5 healthy volunteers with a minimum age of 18 were recruited. The final sample size for the patient cohort was determined following completion of the retrospective pilot study and comprised 72 patients (including 20% drop-out rate) with known or high clinical suspicion of MS and at least 2 identifiable FLAIR-hyperintense lesions. During the second prospective stage, from July 2024 to February 2025, 72 consecutive patients with known or high clinical suspicion of MS and at least 2 visible FLAIR-hyperintense lesions (40 women; mean age, 40.1 years) underwent MR imaging as indicated by the treating physician, at the Institute of Radiology and Neuroradiology, Evangelisches Krankenhaus Oldenburg, using a 1.5 T MR scanner. General exclusion criteria included: non-MR-conditional implants such as pacemakers or automatic defibrillators, implanted neural stimulators, ocular metallic foreign bodies, devices requiring a safety mode that poses increased risk to the patient (e.g., temporary deactivation of defibrillator function in MR-conditional devices), conditions that could impede study participation (e.g., morbid obesity, severe claustrophobia) and/or unnecessary or avoidable risks such as pregnancy, recent surgery of uncertain stability, or other individual health factors or age-related concerns making MRI burdensome. Specific exclusion criteria for the preliminary study were known concurrent severe central nervous system disorders, except for MS.

**IMAGE ANALYSIS**

For each FLAIR acquisition, regions of interest (ROIs) were drawn on transversely reformatted slices to calculate SNR and CNR within the following regions: 1. an MS lesion, 2. normal-appearing white matter in three predefined locations (two in the supratentorial white matter and one in the brainstem), and 3. background noise. Care was taken to avoid lesion borders. The SNR was calculated as follows: SNR = (mean signal intensity of GRAY MATTER) / (standard deviation of AIR). The contrast-to-noise ratio (CNR) was calculated as follows: CNR = (mean signal intensity of LESION − mean signal intensity of WHITE MATTER) / (standard deviation of AIR).

**SNR, CNR AND BMI**

There was no significant correlation between BMI and SNR for either of the two acceleration methods. This was also true between BMI and CNR for GRAPPA. In contrast there was a weak, but significantly positive correlation between the BMI and CNR in CS (Spearman r = 0,311; p = 0,010). The slight positive correlation between BMI and CNR observed in CS acquisitions may relate to the nonlinear reconstruction properties of CS. CS reconstruction uses sparsity constraints and regularization, which can suppress noise and alter image contrast. As a result, differences in baseline signal characteristics, potentially influenced by coil loading and RF field distribution in larger body habitus, may affect CNR after CS reconstruction. However, this interpretation remains speculative and warrants further investigation.

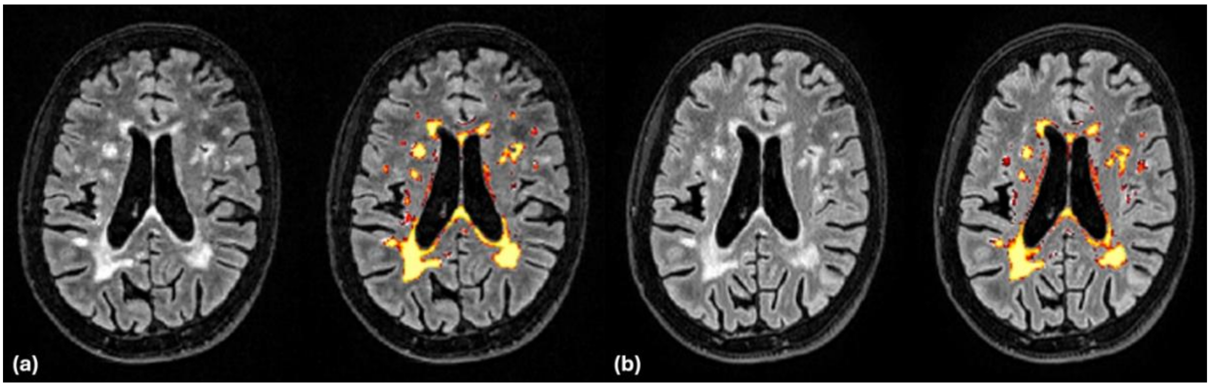


**Figure S1.** Comparative visual performance of automated lesion volumetry for (A) CS- and (B) GRAPPA-accelerated 3D-FLAIR sequences in axial reformats above the level of the basal ganglia.
